# Developing and Evaluating an Online Educational Program for Falls Prevention Care in Community Optometric Primary Care Settings: A Pilot Study

**DOI:** 10.64898/2026.06.22.26355920

**Authors:** Si Ye Lee, Khyber Alam, Jason Charng, Hamed Niyazmand, Jacqueline Francis-Coad, Anne-Marie Hill

## Abstract

**Introduction:** Globally, falls are the leading cause of injury hospitalisation, with vision being a significant falls risk factor. Community optometrists, as primary eye care professionals, are well positioned to contribute to falls prevention care. However, scant studies have evaluated if education could enable optometrists to incorporate falls prevention care into practice. This two-phase pilot study aimed to design and develop an online education program for community optometrists to deliver primary falls prevention care and to evaluate optometrists’ reaction to, and learning from, the education.

**Methods:** In phase one, an education program was designed by optometrists and falls experts and published online. In phase two, community optometrists were recruited through convenience sampling to undertake the education. Guided by the New World Kirkpatrick model® of training evaluation, reaction and learning were evaluated using pre/post surveys. Quantitative data were analysed using Wilcoxon sign-rank tests and McNemar Exact Tests and qualitative responses using inductive content analysis.

**Results:** Participants (n=13) reported high levels of satisfaction and engagement with the online education and unanimously endorsed its relevance to clinical practice. Participants demonstrated significantly improved knowledge and awareness of falls prevention post-education, compared to pre-education and were significantly more confident to enact falls prevention care. Perceived enablers to providing falls prevention care included having access to practical resources and ongoing education. Time constraints during consultation and cost to patients for further care if subsequent referrals were made were identified as possible barriers to providing falls prevention care.

**Conclusion:** Online education improved community optometrists’ knowledge and confidence to provide falls prevention care. Further research that evaluates the effectiveness of continuing education for optometrists to enact falls prevention care into practice is required.

## Introduction

Falls are a global healthcare and societal challenge, with approximately 30% of community-dwelling older adults (aged 65 and above) experiencing a fall annually.(1, 2) Globally, accidental falls are a leading cause of injury hospitalisation and death.(3) In 2023-24, the direct healthcare costs of falls in Australia were estimated to be over $5 billion AUD (2.6 billion GBP).(4) Falls are frequently due to multifactorial risk factors (5) and appropriate treatment can be facilitated through a multidisciplinary team.(6) There is strong evidence that effective multidisciplinary interventions to reduce falls among community-dwelling older adults incorporate a vision component.(6, 7) Consequently, community optometrists are well positioned to strongly contribute to multidisciplinary falls prevention efforts.(8)

Optometry Australia is the professional peak body representing Australia’s registered optometrists. Optometry Australia’s falls prevention guidelines include recommendations for addressing falls risk by tailored prescription management, adapting case histories to assess falls risk, and referring patients to allied health professionals for home modifications.(9) Recently published Australian government falls guidelines (10) support the principles of Optometry Australia guidelines (9) and contain additional comprehensive recommendations that accord with the World Falls Guidelines (WFG).(6) Recommendations include that all health professionals undertake opportunistic case finding, falls risk screening, assessment, and appropriate management that includes providing tailored interventions, shared decision making, and patient education. Specific vision recommendations are to provide early referrals for cataract surgery, prescribe appropriate spectacle lenses for older adults and provide advice to older adults about safe mobility when there are prescription changes.(10)

In Australia, Medicare provides an annual rebate that allows older adults to receive optometry services to assess and manage their eye health,(11) hence optometrists regularly assess and manage older adults in their clinical practice. As they age, adults have an increased risk of chronic eye conditions (12) and visual impairments, such as cataracts, age-related macular degeneration, glaucoma and refractive errors.(13) The resulting decline in visual function due to these impairments is strongly associated with an increased risk and incidence of falls.(14) A recent narrative review highlighted visual acuity, contrast sensitivity, stereoacuity, and visual fields as key ocular assessments that should be conducted in clinical practice to determine falls risk.(15) Large, multifactorial, randomised trials have found that vision interventions when combined with other interventions effectively reduce falls.(16) Randomised trials of individual vision interventions have demonstrated that providing single vision lenses for active, older adults and early cataract surgery for older adults reduces falls.(17, 18)

However, despite the strong association between visual impairments and falls and evidence for the efficacy of vision interventions to reduce falls, there is scant research that has evaluated if optometrists are implementing current recommendations for falls prevention care.(7, 19) A recent scoping review found that community eye care practitioners have low levels of awareness about falls, and may not be conducting falls screening or targeted ocular assessments for falls risk.(7) Research has highlighted the need for community optometrists to improve their translation of evidence-based falls prevention into clinical practice.(20) Theories of behaviour change explain that awareness, knowledge, goals, social influences and self-efficacy influence health professionals’ behaviour in professional practice.(21) Optometrists in Australia are required to complete Continuing Professional Development (CPD) hours as part of their professional registration. However, while optometrists have systematic exposure to CPD topics specific to ocular health assessment and management, there is scant optometry CPD that is specific for falls prevention care.(22, 23)

Implementation science frameworks explain that gaps in training and education are barriers to implementation of evidence-based knowledge into clinical practice.(24) Education plays a vital role in improving health professionals’ knowledge, skills, attitudes, and behaviour related to providing evidence-based healthcare.(25) Evidence from studies of health professionals’ practice suggests education can strengthen the capability, confidence and motivation to deliver falls prevention care.(26) This impact is evident when educational interventions are interactive, multifaceted, and embedded within clinical practice.(27) In the context of falls prevention, structured education through coaching or interactive workshops has led to improvements in allied health professionals’ ability to detect falls risk factors, apply evidence-based interventions, and incorporate falls screening.(28) However, optometrists unlike other allied health professionals, have limited access to education specifically focused on falls prevention care.(19, 23) Online-based education can provide health professionals with increased access to training, support flexible learning, and enable rapid dissemination of evidence-based practice (29). Therefore, the research team planned to develop an online CPD education program to increase optometrists’ capability (knowledge and awareness) and confidence to deliver evidence-based falls prevention care. However, prior to conducting a large implementation study to evaluate the education program, it was important to conduct a pilot study that enrolled community optometrists to identify any potential refinements to the program.(30)

The objectives of the pilot study were to: i) design and develop an online educational program specifically for community optometrists regarding delivering falls prevention care for community-dwelling older adults; ii) evaluate optometrists’ reaction to the online education; and iii) evaluate the efficacy of the online education on optometrists’ learning (knowledge, awareness, and confidence) about falls prevention care.

## Materials and Methods

### Design

A two-phase pilot study was conducted *(see Figure 1*). This allowed the research team to test the feasibility of the efficacy of the online education, identifying potential enablers and barriers to delivering and evaluating the online education program before a larger study was conducted, and to gain valuable insights on how community optometrists reacted to the online education program.(31) The study was undertaken from October 2025 to February 2026. The design was guided by factors for developing e-learning in healthcare (29) (*see Supplementary Table 1*) and adult learning principles (32) (*see Supplementary Table 2*). The Guidance for Reporting Involvement of Patients and the Public (GRIPP2-SF) guideline, which provides a checklist for reporting public involvement in health and social care research, informed the reporting of the study (33) (*see Supplementary Table 3*).

**Figure 1.**
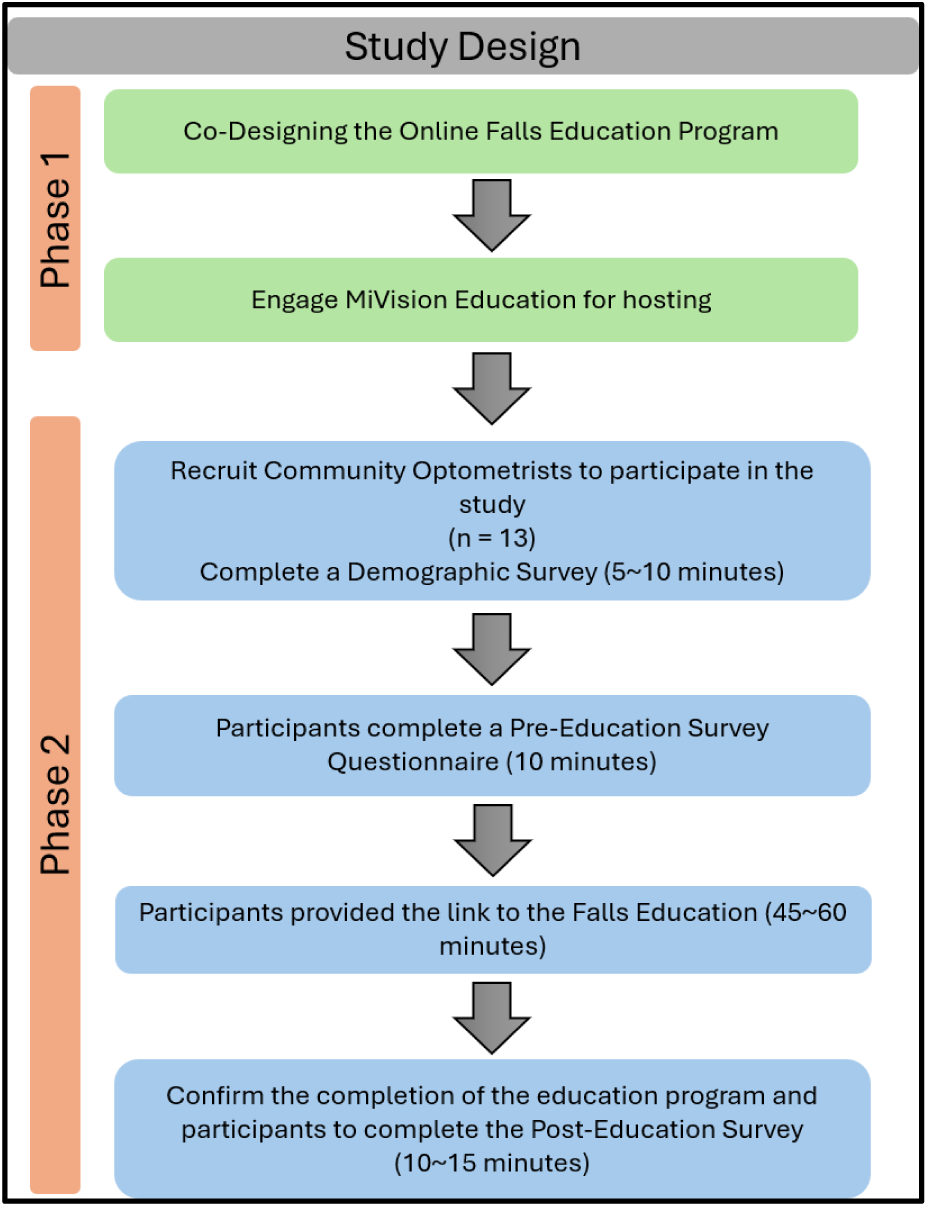
Study Design

### Ethics

This study received ethics approval from the Human Research Ethics Committee at The University of Western Australia HREC (2025/ET000729). All participants received verbal and written information about the study and provided written, informed consent to participate.

### Setting

The education program was delivered via MiVision online platform.(22) MiVision Education is an independent, global, public clinical learning platform that launched in 2011 for the optical industry and offers ongoing accredited education for eye care professionals. The platform publishes courses and modules developed by eye care professionals, including opticians, optometrists and ophthalmologists, contributing to their CPD.

### Participants and Recruitment

Following guidelines for a pilot study, the sample size was based on evaluating the feasibility and protocols for the larger planned study.(31) A convenience sample of approximately 10 optometrists was planned to gain feedback about the online education program and evaluate knowledge changes pre/post education.(34)

To meet inclusion criteria, participants were required to be an optometrist currently practicing in an Australian community setting, a registered health professional, and treating older adults (≥ 65 years) in their clinic. The study excluded optometrists who worked solely in hospital settings, ophthalmology clinics, or residential aged care. There were no restrictions regarding years of practice or weekly practice hours.

### Phase One: Design of the Education

The education was co-designed by the research team. Four team members were qualified optometrists, three of whom practiced regularly in a clinical setting, and thereby had relevant perspectives about the design and content of education program for community optometrists.

These four members also had educational experience in training postgraduate optometrists, with one member being an education curriculum expert in university optometric courses. The remaining two members of the team were registered physiotherapists who had clinical experience working with older adults in falls prevention, formal educational qualifications, and extensive experience in research that focused on designing, delivering and evaluating falls education.(35, 36) The education design and content was informed by prior systematic and scoping reviews and a focus group study that interviewed clinical optometrists regarding their knowledge about falls prevention practice.(7, 20, 37) Aligning with other CPD education on the MiVision Education platform,(22) the education program was designed to last approximately 45 to 60 minutes. The content was presented using short videos, images, texts and figures that presented summative information, based on falls prevention guidelines, about the epidemiology of falls, risk factors for falls and relevant treatment options tailored for community optometrists.(6, 9, 10) The online education program provided key strategies to guide optometrists regarding the concepts of delivering falls prevention care for older adults and aimed to facilitate subsequent behaviour change in clinical practice.(38) Literature suggests online-based education can have similar effectiveness in improving knowledge compared to traditional methods of education.(29)

The education program learning objectives for participants were to; i) identify the impact of falls on older adults in the community; ii) understand the strong associations between age-related vision changes, ocular diseases and falls; iii) understand the key elements for embedding falls screening, assessment, treatment and referral into optometric practice; iv) improve awareness of the recommended adaptations and modifications for glasses prescriptions for older adults who have a history of falling; v) improve confidence to discuss falls prevention with older adults as part of optometric practice; vi) identify relevant falls guidelines that are specific for optometric practice; and vii) identify available falls prevention resources to use in clinical practice.

The education provided free e-resources for the participants to access on education program completion, which aligned with best practice for health professional education.(39) This aimed to provide further opportunity for independent learning and assist participants to build confidence and enact learning into practice.(40) Free access to falls prevention handouts for older adult patients, falls prevention guidelines and education resources on general aspects of falls prevention for health professionals were provided through online links. Participants received information on how to join a falls community of practice for health professionals in Western Australia for ongoing peer learning and practice support.

### Phase 2: Evaluation of the Education

The outcomes evaluated were based on the New World Kirkpatrick Model® for evaluating training programs.(41) This established framework provides a systematic approach for evaluating the effectiveness of training and education at four hierarchical levels; i) reaction - initial learner reaction to the education; ii) learning - learner acquiring the intended knowledge through engagement with the education; iii) behaviour - measuring how learners applied the knowledge obtained during the education; iv) impact - impacts of the learning on desired behavioural outcomes.(41) For this pilot study, the first two levels of the framework, reaction and learning, were used to systematically evaluate the education program. The outcomes measured specific to each level are presented in *Table 1*.

**Table 1:** Kirkpatrick framework for evaluating the education(41, 42)

| **Level of Kirkpatrick Model** | **Outcomes Evaluated** |
| --- | --- |
| Level 1: Reaction<br>(*measured post-education*) | • Participants' satisfaction with the education program<br>• Participants' engagement with the education program<br>• Participants' perceptions about the relevance of the education program to their clinical practice |
| Level 2: Learning<br>(*measured pre /post-education*) | • Changes in participants' knowledge and awareness of falls prevention care in optometric practice<br>• Changes in participants' attitudes towards incorporating falls prevention care into their clinical practice<br>• Participants' confidence to incorporate falls prevention care into their clinical practice |

Reaction evaluated the participants’ satisfaction and engagement with the content and delivery of the education, and their perceptions of the relevance of the education to their clinical practice. Participants were asked to provide open comments offering any suggested improvements for the education program. Learning evaluated changes in participants’ knowledge and awareness regarding falls prevention care, and attitudes and confidence towards falls prevention treatment (41, 42). Participants were asked to identify any perceived barriers or enablers to incorporating the information provided in the education program into their clinical practice. Each outcome was measured using the appropriate multiple-choice, Likert-scale, dichotomous, and open-ended questions. The pre-/post-education surveys are presented in full in *Supplementary Table 4*.

### Phase 2: Data Collection and Procedure

The education program was published online through the MiVision Education platform.(22) For purposes of the pilot study, the education was made available through password-protected viewing only to the participants enrolled in the study. After providing informed consent, participants completed a demographic survey (age, university qualifications, employment arrangements, approximate number of patients treated weekly, proportion of older adults treated and any previous training in geriatric eye care, falls prevention or low vision training). A pre-education survey was then provided to participants which evaluated their baseline knowledge, awareness, and confidence regarding the topic of falls and falls prevention treatment (*see Table 1*). Upon completion of the pre-education survey, an online link to the falls prevention education was made available to the participants. Given the time constraints faced by clinical optometrists, participants were given two weeks to complete the education. Completion of the education triggered automatic access to the e-resources.

After two weeks, the research team followed up with participants to confirm completion of the education program. Subsequently, participants were provided with the post-education survey. All surveys were administered through a secure online RedCap portal held at the University.(43)

## Data Analysis

### Quantitative data

All survey data were de-identified and analysed using SPSS software (IBM, Version 30, 2024). Quantitative data, including participants’ demographic profile, were summarised using descriptive statistics (frequency, percentages, mean, standard deviation (SD) or median, Inter Quartile Range (IQR) for non-parametric data). Participants’ levels of satisfaction, engagement and perceived relevance of the education were evaluated in post-education surveys utilising Likert scale and dichotomous items. Changes in participants’ knowledge, awareness, attitudes and confidence regarding falls prevention were assessed by comparing the pre and post-education survey responses, with the analysis determined by the type of survey data collected. For Likert-scale measures and individual ordinal Likert items, such as self-rated confidence and attitudes, the Wilcoxon signed rank test was employed to assess within-participant changes. Binary survey items were analysed with McNemar’s test to assess changes in paired categorical responses.

### Qualitative data

All open-ended responses were transcribed verbatim into Microsoft Excel (2010) and imported into NVivo (Lumivero, US, Ver. 14, 2023) for coding. Two researchers (SYL and KA) independently read through all responses several times to familiarise themselves with the data. The open-ended survey responses were subjected to inductive content analysis, with codes developed iteratively and grouped into emergent themes.(44, 45) Both researchers met frequently to discuss and compare their coding, and any disagreements were resolved through discussion with a third independent senior researcher (AMH). The confirmability of the qualitative findings was ensured by using verbatim participant quotes.(46) All participant quotes were de-identified and represented using pseudonyms to maintain confidentiality.

## Results

### Participants’ Characteristics

Thirteen participants (n = 11 (84.6%) female, n = 2 (15.4%) male) enrolled in the study, demographic characteristics are presented in Table 2. Participants reported treating an average of 46 patients per week. Nine (69.2%) participants reported they had not completed any low vision or falls prevention training, while four (30.8%) participants responded that they had completed training in managing low vision, but not falls prevention.

**Table 2:** Participants’ Characteristics.

| Characteristics<sup>a</sup> | Participants n = 13 (100%) |
| --- | --- |
| Age |  |
| 20 to 29 | 9 (69.2%) |
| 30 to 39 | 1 (7.7%) |
| 40 and above | 3 (23.1%) |
| Gender |  |
| Female | 11 (84.6%) |
| Male | 2 (15.4%) |
| State of Qualification |  |
| Victoria | 8 (61.5%) |
| South Australia | 1 (7.7%) |
| Queensland | 1 (7.7%) |
| New South Wales | 1 (7.7%) |
| Western Australia | 2 (15.4%) |
| Location of work |  |
| Queensland | 2 (15.4%) |
| Victoria | 4 (30.8%) |
| Western Australia | 7 (53.8%) |
| Employment |  |
| Independent community practice (*full-time or part-time*) | 1 (7.7%) |
| Corporate community practice (*full-time or part-time*) | 7 (53.8%) |
| Others | 5 (38.5%) |
| University clinic | 3 |
| Government funded clinic | 2 |
| Years of experience, mean (SD) | 8.6 (8.3) |
| Number of patients seen weekly, mean (SD) | 45.6 (27.5) |
| Proportion of older adults seen |  |
| < 25% | 2 (15.4%) |
| 25 – 50% | 6 (46.1%) |
| 51 – 75% | 4 (30.8%) |
| > 75% | 1 (7.7%) |
| Low vision or falls prevention training |  |
| No | 9 (69.2%) |
| Yes | 4 (30.8%) |
| Low vision training during university course | 3 |
| Low vision training during further training | 1 |

<sup>a</sup>All data measured in n (%) unless stated otherwise

## Reaction to the education

### Satisfaction

Participants reported highly positive reactions to the education format and delivery. The online program was rated as easily accessible [n = 7 (53.8%) strongly agree; n = 6 (46.2%) agree], with no participants reporting any difficulties with accessing the education through the MiVision platform. The average time to complete the education was 26.2 (SD 14.2) minutes. All participants (100%) were satisfied that the purpose of the education was described clearly, and twelve (92.3%) were satisfied that the sequence of the content was logical, with one participant disagreeing with the presentation sequence. Participant P6 commented, “*The education program was easy to follow, and the information was given in an appropriate sequence*”. Eleven (84.6%) participants rated the length of the education as satisfactory. Twelve (92.3%) participants agreed that the program satisfactorily met their learning needs.

### Engagement

Eleven (84.6%) participants rated the range of multimedia content as appropriate. The two short introductory videos were viewed positively by nine participants (69.2%) who agreed with the video length. All participants agreed that the inclusion of section breaks and summaries made the education easy to digest. “*The CPD was simple to digest and broken down into dot points that are easy to remember for optometrists*” (P2). The education program content was highly rated by all participants, with nine (69.2%) agreeing and four (30.8%) strongly agreeing that they had a positive reaction to this content. The vision impairment images were also positively rated by ten participants; however three participants felt they would prefer more images in the education with one participant stating, “*More picture illustrations would be better*” (P10). One participant suggested that a case study would be beneficial: “*Clinical scenarios or case studies to simulate practical scenarios*” (P3).

### Relevance

All participants (n = 8 strongly agree, n = 5 agree) endorsed the falls prevention education being relevant for their practice after completing the education. Compared to pre-education ratings where three participants strongly agreed, nine participants agreeing and one participant being undecided (pre-education median 3.0 [IQR 3–3.5], post-education median 4.0 [IQR 3–4], Z = - 2.121, p =.034). Eleven (84.6%) participants agreed that the education provided clear instructions for incorporating falls prevention care. “*The advice on lighting conditions and the discussion around early cataract surgery has been very helpful with how I will discuss cataracts with my patients and encouraging patients not to delay cataract surgery*” (P7). Ten (76.9%) participants agreed the education offered manageable actions that could be applied in practice.

“*This* [education] *has encouraged me to make sure that I screen my elderly patients better and provide them with the support they need day to day*” (P9).

### Changes in knowledge and awareness about falls prevention care in optometric practice Knowledge

Pre/Post changes in participants’ knowledge about falls prevention care pre and post-education are presented in *Figure 2, Tables 3 and 4 and Supplementary Tables 5 and 6*.

**Figure 2.**
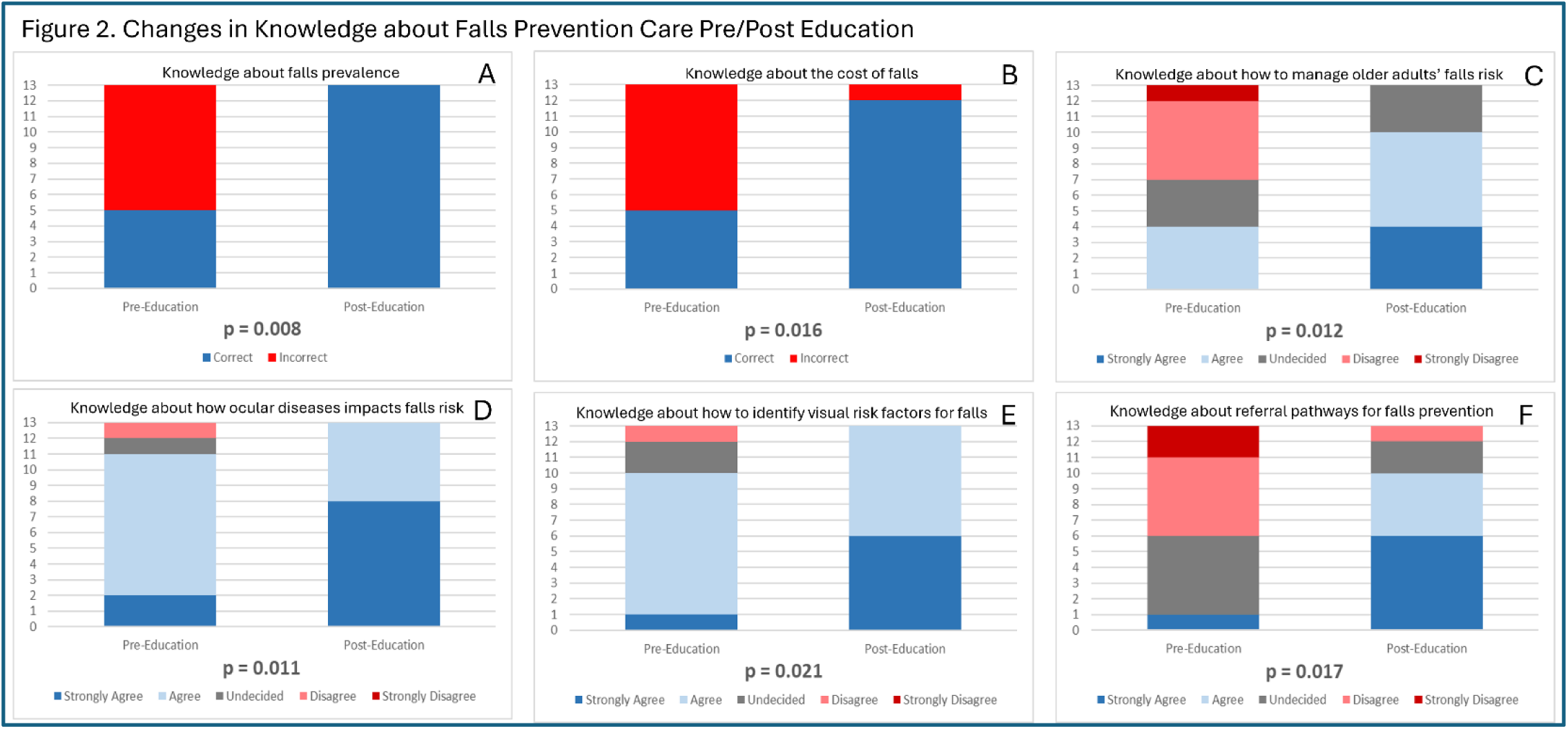
Changes in Knowledge about Falls Prevention Care Pre/Post Education

**Table 3:** Comparisons of Pre/Post knowledge, awareness and confidence about falls prevention care.

|  | Knowledge |  |  |  | Attitudes |  |  |
| --- | --- | --- | --- | --- | --- | --- | --- |
|  | Knowledge to manage older adults' falls risk | Knowledge of ocular diseases and falls risk | Knowledge to identify visual risk factors for falls | Knowledge of referral pathways for falls prevention | Relevance of Falls Prevention Care in Optometry | Willingness to Incorporate Falls Prevention Care | Willing to initiate referral to allied health professionals |
| Pre-Education Median (IQR) | 2.0 (1 – 3) | 3.0 (3- 3) | 3.0 (2.5 – 3) | 1.0 (1 – 2) | 3.0 (3 – 3) | 3.0 (3 – 4) | 3.0 (3 – 4) |
| Post-Education Median (IQR) | 3.0 (2.5 – 4) | 4.0 (3 – 4) | 3.0 (3 – 4) | 3.0 (2.5 – 4) | 4.0 (3 – 4) | 4.0 (3 – 4) | 4.0 (3 – 4) |
| Z-Scores | -2.520 | -2.530 | -2.309 | -2.379 | -2.111 | -1.633 | -2.236 |
| Asymp. Sig. (2-tailed) | .012* | .011* | .021* | .017* | .035* | .102 | .025* |
| Note: Comparisons analysed using Wilcoxon Signed Rank Test<br>*Statistically significant |  |  |  |  |  |  |  |

**Table 4.** Comparisons of pre/post levels of knowledge about falls epidemiology.

|  | Knowledge of Falls Prevalence<sup>a</sup> | Knowledge of the cost of falls<sup>a</sup> | Awareness of falls prevention guidelines<sup>b</sup> | Awareness of falls prevention resources<sup>b</sup> |
| --- | --- | --- | --- | --- |
| Pre-education | 5/8 | 5/8 | 2/11 | 4/9 |
| Post-education | 12/1 | 13/0 | 13/0 | 9/4 |
| Exact Sig. (2-tailed) | .016* | .008* | <.001* | .063 |
| Note: Comparisons analysed using McNemar's Exact Test |  |  |  |  |
| *Statistically significant |  |  |  |  |
| <sup>a</sup> Results presented as correct/incorrect |  |  |  |  |
| <sup>b</sup> Results presented as yes/no |  |  |  |  |

Participants provided significantly more correct responses to falls epidemiology questions (*see Figures 2A and 2B*) after completing the education. Participants rated their levels of knowledge about how to manage older adults’ falls risk as significantly higher after completing the education (*see Figure 2C*). Participants were significantly more knowledgeable about the association between ocular diseases and risk of falls and which deficits in visual function (e.g. contrast sensitivity) are associated with increased falls risk after completing the education (*see Figures 2D and* 2E). Finally, participants were significantly more knowledgeable about what referral pathways were available for older adults to receive multidisciplinary falls prevention care after the education (*see Figure 2F*).

Participants also demonstrated knowledge gain after the education in their clinical knowledge about falls prevention care (*see full responses in Supplementary Table 6*). Screening questions participants used to determine patients’ falls risk became more focused and aligned with guideline recommended practice, moving from broader generic questions, for example “*How do you go with navigating your environment day to day?*” (P9, pre-education) to specific questions “*Have you had any previous falls in the last year? Any concerns about falls or limited mobility?*” (P9, post-education). Participants were initially able to identify some visual assessments for determining falls risk, such as “*Visual acuity and visual fields*” (P4, pre-education). After completing the education, they were able to describe a broader, evidence-based falls risk assessment including, “*Visual acuity, contrast sensitivity, visual fields and stereoacuity”* (P4, post-education). Considerations of falls risk were evident in changes in participants’ descriptions of optical treatment. One participant commented (on their former treatment) “*Choose appropriate size frames, avoid multifocal/bifocal in new elderly wearers*” (P12), but after the education provided more evidence-based treatment options: “*Avoid large changes to refraction, avoid multifocal or bifocal in elderly patients who have not worn them before, for existing multifocal or bifocal wearers consider single vision distance spectacles for use outside the home*” (P12). Similarly, another participant’s response describing an optometrist’s overall strategy for falls prevention care expanded from a single strategy, such as “*Ask about prior falls or balance issues*” (P7, pre-education) to more comprehensive care “*Consider single vision instead of bifocals or multifocal, ask in history about falls/risks, correct distance refractive error, discuss falls risks with patients, initiate referrals to other practitioners involved in care*” (P7, post-education).

### Awareness

Changes in participants’ awareness of falls prevention care pre/post-education are presented in *Figure 3, Table 4, and Supplementary Table 5*.

**Figure 3.**
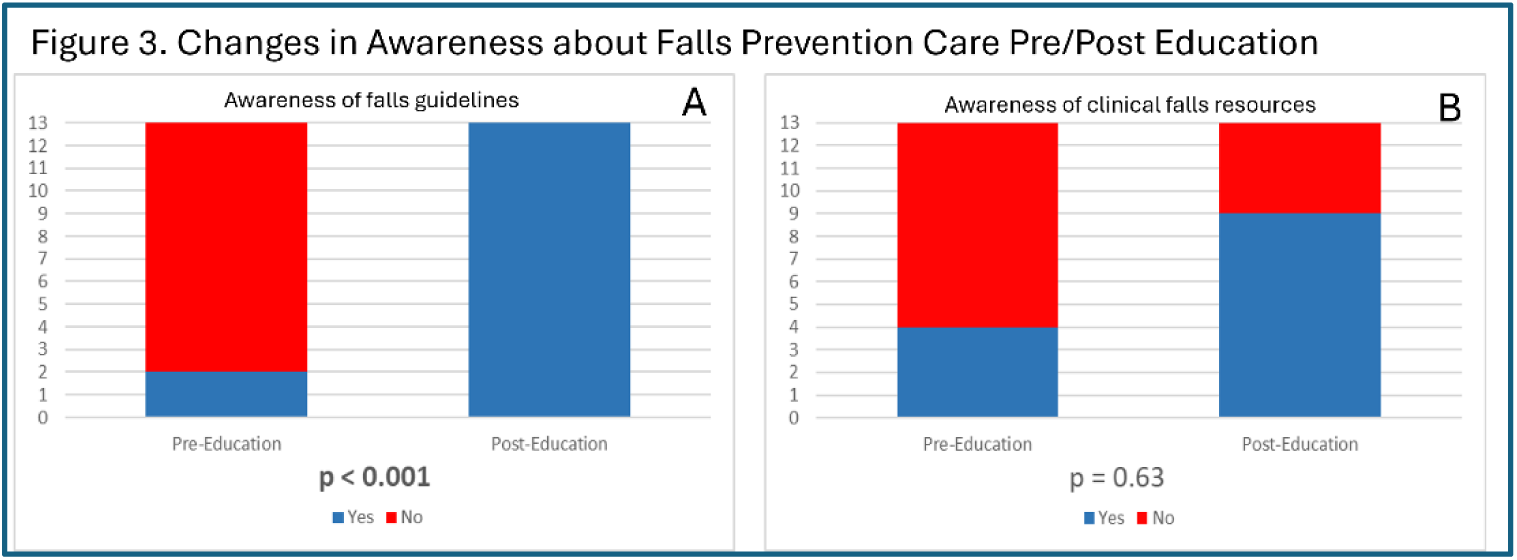
Changes in Awareness about Falls Prevention Care Pre/Post Education

Compared to baseline, participants were significantly more aware of the availability of falls guidelines for use in clinical practice after completing the education (*see Figure 3A*). This improvement was exemplified by one participant’s comment: “*I am more informed on the general* [falls] *guidelines and resources available to educate myself and my patients*” (P3). Participants also demonstrated improved awareness of the falls prevention resources available for staff and patients, but this improvement was not significant (*see Figure 3B*). One participant commented, “[The education] *made me more aware of what to ask patients, and now I know where to find resources*” (P4).

### Changes in attitudes towards incorporating falls prevention care in optometric practice

Participants’ changes in attitudes about falls prevention care pre and post education are presented in *Figure 4, Table 3 and Supplementary Table 5*.

**Figure 4.**
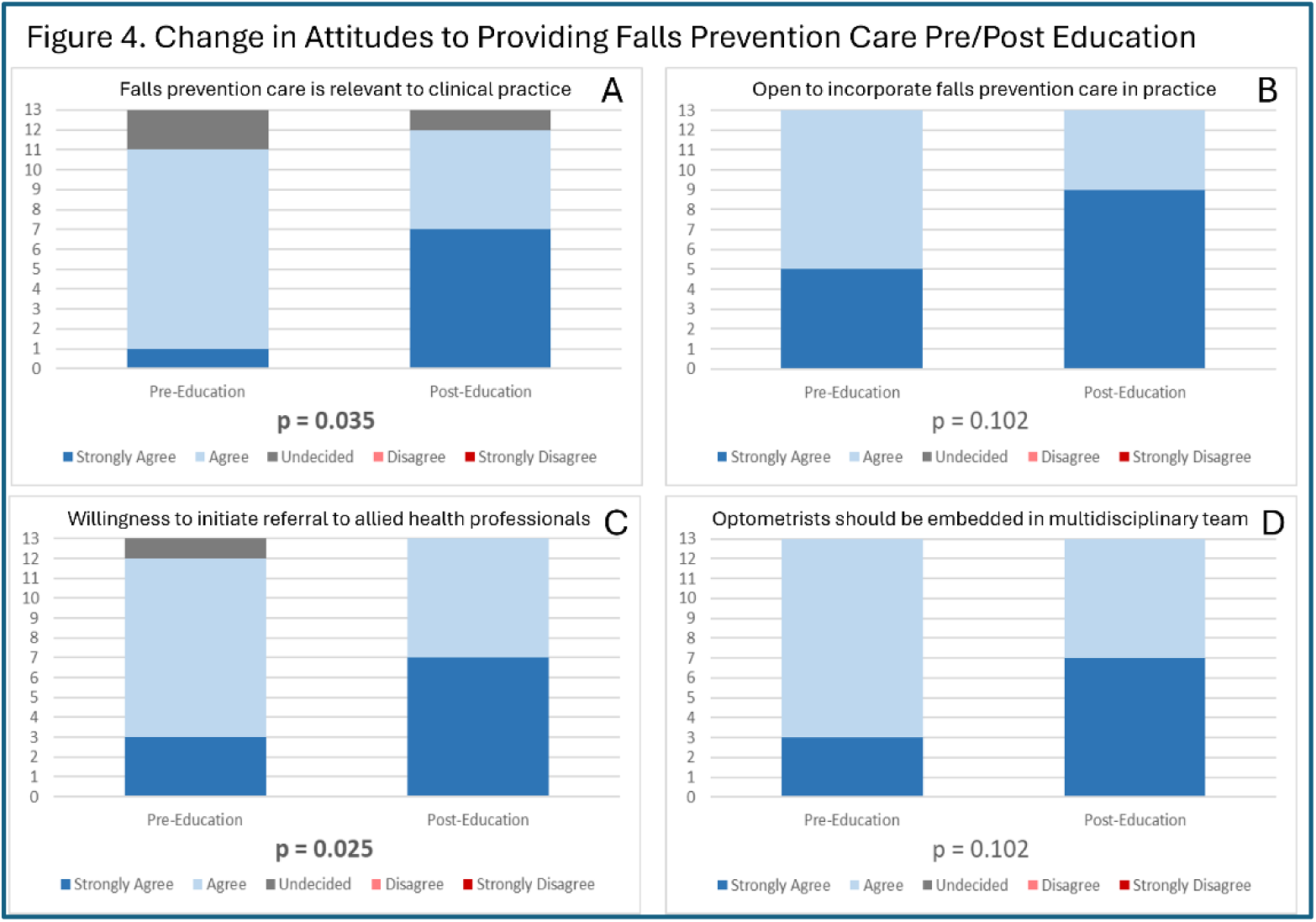
Changes in Attitudes to Providing Falls Prevention Care Pre/Post Education

Compared to baseline, participants showed significantly more positive attitudes regarding the relevance of falls prevention to their clinical practice after the education (*see Figure 4A*). They were more willing to incorporate falls prevention when providing care to older adults (*see Figure 4B*), “*I have learnt about the implications* [of falls risk] *and I am more motivated to manage fall risk*” (P6). All participants (n = 7 strongly agree, n = 6 agree) were significantly more willing to initiate referrals to other allied health professionals after the education (*see Figure 4C*, 3 = strongly agree, 10 = agree, 1 = undecided). Similarly, participants more strongly agreed that optometrists should be part of multidisciplinary teams for falls prevention care compared to pre-education (*see Figure 4D*). This was supported by a participant commenting, “*Yes, it has reminded me that we play a useful role in* [falls prevention]” (P13).

### Changes in confidence to incorporate falls prevention care in optometric practice

Compared to baseline, participants felt significantly more confident to provide appropriate falls prevention care to older adults during clinical practice [pre-education median 2.0 (IQR 1–2), post-education median 3.0 (IQR 3–4), Z =-2.86, p =.004]. One participant reflected, “[The education] *makes me more confident in managing older adults’ falls risk. The statistics also help in linking* [older adults’] *ocular findings and fall risks*” (P5). Participants’ described how the education had improved their confidence in knowing what specific strategies to perform, “*I now feel more knowledgeable and confident in my ability to screen for falls risk and be able to take actionable steps to reduce the risk of falls in my patients*” (P1) (*see Supplementary Table 5*).

### Barriers and enablers for optometrists incorporating falls prevention care Barriers

Participants perceived several barriers to incorporating falls prevention care into optometry practice (*see Supplementary Table 5*), with time constraints emerging as a common concern. They believed that adding falls prevention assessments could extend consultation times, making it challenging to complete all the required optometric clinical assessments. This perception appeared to be more evident among participants working in fast-paced settings with shorter consultation times. One participant explained “[For] *optometrists working in retail practices (20-30 min appointments) there may not be enough time to ask all questions and do all the appropriate tests*” (P4).

Some participants believed patient level barriers could impact their delivery of falls prevention messages, such as older patients having limited awareness of falls risks and being reluctant to seek help due to fears about losing independence. As one participant explained, “*Patient lack of awareness and fear of losing independence*” (P3).

Further perceived barriers were related to system level challenges that could make it difficult for optometrists to refer patients for appropriate falls prevention treatment. Examples provided were, public hospital eligibility criteria for cataract surgery and the cost or availability of other allied health services, such as an occupational therapy home visit. Financial barriers were perceived as particularly important for patients requiring an ophthalmology referral. “*Cost is a barrier when it comes to being referred for cataract surgery, ongoing glaucoma or macular degeneration management as those in* [regional area] *there is no public ophthalmology care nearby*” (P5). Participants further highlighted equipment related challenges, suggesting that tools such as the Melbourne Edge Test could support routine assessments if they were more readily available “*Ready availability of the Melbourne Edge Test (small, easy and portable)*” (P11).

### Enablers

Participants suggested enablers that could support optometrists in incorporating falls prevention care into practice (*see Supplementary Table 5*). They emphasised that providing practical clinical falls prevention resources, including infographics, screening tools, and educational materials could support optometrists’ practice. Utilising concise, accessible guidance was seen as particularly helpful, with one participant suggesting “*A page printout or webpage that can be saved as a tab on the computer in dot point guideline form would be really good to have as a reference to refer to during consult*” (P2). Others highlighted the importance of education and training, especially ongoing professional development to ensure all staff were confident to implement falls prevention care “*Ongoing staff education so other practitioners are well informed of the available falls prevention guidelines*” (P3). Participants also described the value of multidisciplinary collaboration, expressing a need for improved optometrist awareness of the role of general practitioners and allied health professionals in preventing falls. This was exemplified by one participant, who stated “*Awareness of other multidisciplinary services and referral pathways for patients to access physiotherapy, podiatry etc*” (P12).

## Discussion

Community optometrists who completed a tailored, online falls education program reported high levels of satisfaction and strongly endorsed the relevance of the program. Participants showed significant improvements in knowledge, awareness, attitudes and confidence to undertake falls prevention care during clinical practice after receiving the falls prevention education, compared to prior to the education. Some participants were uncertain about the relevance of falls prevention care for optometrists prior to completing the education, but after completion strongly agreed that falls prevention care was relevant for their clinical practice. Behaviour change theory explains that improving capability (which includes both knowledge and awareness) is an essential component for facilitating behaviour change.(21)

The online education was tailored specifically for optometrists and provided relevant information based on their clinical practice. This aligns with adult learning theory, which explains that adult learners seek to identify the relevance of the learning and apply knowledge gained to solve real life problems.(32) The principles of online education and health professional education further supported the creation of the falls prevention education.(29, 47) It was feasible to deliver the education online because falls prevention is a novel topic for optometrist working in primary care (37) and optometrists regularly engage in online CPD sessions to be appraised of recent evidence based practice. Hence the online education format is familiar to optometrists.(22, 48) Previous falls programs have been designed for specific health professional groups such as occupational therapists or physiotherapists (25, 26, 49) suggesting that optometrists, like other health professionals, would benefit from tailored falls education. While hospital or aged care falls prevention is often led by a multidisciplinary team, community optometrists operate as sole primary care practitioners and therefore need education that is relevant and tailored for this context. Community optometrists as primary care professionals can undertake initial screening as recommended by the WFG (6) and provide evidence-based strategies targeting visual risk factors for falls. Optometrists may also be the first point of healthcare contact for an older person to discuss falls,(6) hence it is important that optometrists are able to provide appropriate advice and referrals.

The online education program generated positive reactions, with participants consistently reporting high levels of satisfaction with the accessibility, purpose and sequence of the education. The New World Kirkpatrick Model® explains that positive reactions are fundamental to learners achieving the desired learning outcomes and subsequently enacting the desired behaviours.(42) Participants also responded positively to the range of multimedia used, use of section breaks and summaries to make the education easy to follow. These strategies aimed to ensure the education captured learners’ attention and facilitated a smooth learning experience.(50, 51) Relevance was strongly endorsed with all participants reporting that the education was relevant for their clinical practice. Kirkpatrick’s training model describes that perceived relevance is critical to evaluating reactions to education, because a belief that the information is relevant strengthens an individual’s intention to apply new knowledge, aiding in translation into behaviour change.(41) Previous research evaluating health professional training suggests that high levels of satisfaction and engagement with the education is associated with improvements in knowledge gain.(51–53)

The Capability-Opportunity-Motivation-Behaviour (COM-B) model of behaviour change offered a useful framework when designing this education.(21) Applying the framework suggested that the online education should provide evidence-based strategies for falls prevention care.(21) The education led to significant improvements in participants’ knowledge and awareness reflecting strong Level 2 Learning outcomes. This knowledge gain has potential to meaningfully improve optometrists’ ability to contribute to falls prevention care. Online education training for health professionals that is evidence-based has been shown to consistently improve health professionals’ self-reported knowledge, skills, and attitudes and supported behaviour change in clinical practice (53). Participants’ knowledge gain regarding guideline aligned screening questions, broader visual assessment considerations and more nuanced refraction management reflects recommendations outlined from the WFG and optometry falls guidelines.(6, 9) Studies have highlighted persistent gaps in optometrists’ awareness of falls prevention guidelines, suggesting that limited exposure can contribute to inconsistent practice.(7, 20) The improvements observed in this pilot study indicate that potentially online, evidence-based education can address these deficits.

Education is an essential step to implementation however, implementation theory suggests that successful uptake of evidence-based care requires multilevel planning targeting both providers and practice settings.(54, 55) Participant improvement in capability through an online falls education program facilitated future engagement in falls prevention care. However, despite positive changes in participants’ knowledge, awareness, attitudes and confidence about falls prevention care, they identified perceived barriers and enablers to incorporating falls prevention care in clinical practice. Potential barriers included time constraints and patients’ lack of awareness about falls prevention care. These barriers reflect similar challenges identified in a recent study of community optometrists, which found that short consultation times and competing clinical priorities limited their opportunity to deliver falls prevention care.(20) System level challenges through cost or access to services particularly in regional areas were also viewed as potentially limiting the referral pathways that optometrists could use to support appropriate care for older adults.(56) Potential enablers identified were use of printout infographic or a webpage to access during clinical practice and further education through CPD sessions targeting falls prevention. This aligns with evidence that concise resources for allied health professionals can aid the integration of evidence-based strategies into clinical practice.(8)

### Strengths and Limitations

This study is one of the first to develop and pilot online education tailored specifically for optometrists to improve their knowledge and awareness regarding falls prevention care. This emerging, important focus on falls education for optometrists has been recognised by the United Kingdom College of Optometrists and the World Falls Guidelines with adjoining education support.(6, 57) Our pilot study utilised optometrists as co-design experts and asked community optometrists to directly evaluate their knowledge gain from the education. A key strength was the feedback gained from this pilot study that will assist to improve the education prior to a larger trial being conducted. Participants reported being easily able to complete the education and provided valuable suggestions for minor improvements, such as improving the clarity of picture design, providing a brief clinical infographic and using a case study. Participants also suggested barriers that are specific to optometrists enacting the education in practice. This feedback can inform the design of future falls prevention education for optometrists, for example by including a section on how to address barriers to enactment of suggested strategies.

Limitations to the study were that it sought a small convenience sample and larger evaluations of the program are required to measure optometrists’ knowledge gain and to understand the impact of the education on optometrists’ practice of falls prevention care. The majority of participants were early career optometrists, typically with 1 – 5 years of clinical experience, which may have influenced the findings. However, the mid to late career optometrists who were enrolled described similar gaps in knowledge and awareness regarding falls prevention care. Due to the pre/post design there was no control group to compare changes in knowledge, awareness and confidence. Although a post-education survey was completed immediately after the education, it is unknown if the learning was retained. The pre/post-education survey did not test skills or behaviours (Level 2/3 of Kirkpatrick’s training model) (41) in this study because the online education provided evidence-based theory and did not include a practical component for the optometrists. The learning objectives of the education were to prepare optometrists to translate learnings acquired into clinical practice, by incorporating what they learnt into their older adult consultations. A further study is underway to assess practical skills and behaviour to evaluate how optometrists enact falls prevention care after receiving online falls education. The study was conducted in Australia, and the education may require modifications for use by optometrists in other countries.

## Conclusion

Optometrists as primary eye care professionals are well positioned to provide falls prevention care to older adults during consultations. This pilot study showed that community optometrists who completed tailored, online falls prevention education significantly improved their capability and confidence to deliver falls prevention care compared to their baseline levels. The online education was well received by the optometrists who provided positive responses regarding the design and clinical relevance and offered valuable suggestions for improvements. Further studies are required to evaluate how community optometrists can improve their knowledge about falls prevention care and incorporate that knowledge gain into clinical practice.

## Acknowledgements

We extend our gratitude to all participating community optometrists for taking the time to complete the falls prevention education program and the accompanying pre/post-education surveys.

## Author Contributions

SYL was primarily responsible for drafting the manuscript with support from AMH and KA. SYL, AMH, and KA were responsible for study conception and design with support from JC, HN and JFC. SYL led data collection and procedures with support from KA and input from JC, and HN. SYL completed the data analysis with support from AMH and KA. All authors critically reviewed the manuscript for its content and approved the final version of the manuscript for submission.

## Data Availability Statement

All the data are contained in the manuscript. The raw data are not available due to ethical considerations regarding preserving the anonymity of the participants. Further enquiries can be directed to the corresponding author.

