## Supplementary File for "Developing and Evaluating an Online Educational Program for Falls Prevention Care in Community Optometric Primary Care Settings: A Pilot Study"

Supplementary Files


| | | |
|---|---|---|
| S. Table 1 | Considerations for Developing Online Education | Pg 2 |
| S. Table 2 | Adult Learning Principles for Education Design | Pg 3 – 4 |
| S. Table 3 | GRIPP2- SF Reporting Checklist | Pg 5 – 6 |
| S. Table 4 | Pre/Post Education Survey | Pg 7 – 9 |
| S. Table 5 | Coding matrix of open ended responses with exemplar quotes | Pg 10 - 11 |
| S. Table 6 | Pre/Post Education responses to clinical knowledge about falls prevention care | Pg 12 - 16 |
| | References | Pg 17 |

S. Table 1. Considerations for Developing Online Education(1)

| | Developing online education | Application to optometrist learning |
|---|---|---|
| 1 | Success in achieving the learning goals | • Provided learners with clear learning outcomes at the start of the education<br>• Clear pre post evaluation to assist in identifying achievements<br>• Learners provided feedback on a survey to reflect on their learning |
| 2 | Satisfaction with the course structure | • The education was held on a widely used optometry CPD platform<br>• Education had introduction videos to describe the relevance of the content for optometrists.<br>• Structured flow of information aligned with optometric clinical practice |
| 3 | Availability of practice-based learning and enjoyment | • Education is held online and is accessible with a link provided to learners<br>• Education free<br>• Platform uptime includes the duration of the study<br>• Online links provided to learners to access further resources from injury matters for clinical use and free to download<br>• Clinical strategies are described to learners for active translation to practice |
| 4 | Effectiveness of the course | • A pre-education survey was completed to understand the baseline knowledge of learners<br>• A post-education survey was administered to evaluate the change in knowledge, confidence and awareness of learners.<br>• Learners had access to both surveys to self-evaluate their own learning |
| 5 | Readability, including accessibility and offline learning | • MiVision assisted the research team in editing and formatting the educational content to ensure it reflects a consistent education standard for optometrists<br>• Education is held on a single webpage that the learners can freely navigate |
| 6 | Engagement with the course, ease of use and navigation | • Hosted by experienced online learning platform that is specifically tailored for eye care professionals<br>• Videos and image simulations provide complementary details with the texts |

S.Table 2. Adult Learning Principles for Education Design

| | Adult Learning Principle | Application to optometrists' learning |
|---|---|---|
| 1 | Adult learners need to know why they need to learn something | • Inform the optometrist about patients' risk of falls<br>• Give statements about the size of the problem<br>• Remind that vision is a key falls risk factor and evidence based interventions use vision treatments<br>• Give confirmation that education is approved by leaders in optometry training at the University |
| 2 | Learners should actively participate in the learning process | • Provide an overview and state the objectives of the education<br>• Extra online resources of current falls prevention guidelines for ongoing self-directed learning<br>• Provide self-directed pre post learning evaluation<br>• Access to free online resources for implementing falls prevention in clinical practice for ongoing self- directed learning |
| 3 | Learning should relate to the relevant prior knowledge of the individual | • Provide information to learners about standardised optometric clinical assessments and treatment that can be implemented for falls prevention<br>• Inform learners about the impact of ocular diseases on falls risk<br>• Provide learners with information on how poor visual function is linked to falls risk<br>• Describe how falls strategies fit to current optical assessments and treatments that are familiar to optometrists |
| 4 | Learning includes active engagement and participation | • Breaking content to small chunks for clear messaging<br>• Videos allow learners to reflect on the epidemiology of falls in the community<br>• Use of 'real world' simulations of visual impairments to aid learners to engage in reflective professional thinking to understand the impact on falls risk<br>• Learners are provided a with a free Community of Practice link to meet other health professionals who are interested in falls prevention |
| 5 | Learning encompasses practical and hands on application | • Key strategies listed for optometrists to enact falls prevention for older adults into their clinical practice.<br>• Provide information on roles of different allied health professionals for falls prevention treatment<br>• Provide online links to patient resources from Injury Matters for use by the optometrist |
| 6 | Learning respects individual learning styles and preferences | • Include a range of education modalities (text, videos and images)<br>• Include image simulations of visual impairment for learners to understand 'real world' impact |

| | | |
|---|---|---|
| | | • Several summary images<br>to provide and overview and concise information to participants |
| 7 | A comfortable and encouraging positive learning environment | • Positive and self-directed learning<br>• Links to further reading available to learners for a holistic understanding of guidelines<br>• Links to reports to understand community impact of falls<br>• Able to be done at home anywhere<br>• No formal test required – formative learning with self-test pre/post education<br>• Certificate available to download after post test to confirm completion of the education |
| Based on Knowles et al, 1972(4) | | |

S. Table 3. GRIPP2 – Short Form Reporting Checklist

"**Developing and Evaluating an Educational Program for Falls Prevention Care in Community-Optometric Care Settings: A Pilot Study**"

| Section and Topic | Item | Reported on Page No. |
|---|---|---|
| 1: Aim<br>*Report the aim of PPI in the study* | The study involved community optometrists in both phases to ensure that the education would be applicable and relevant to public settings (optometrists clinical practices in the community) and the recipients of the education (community optometrists).<br><br>The aims of this study were:<br><br>1. Design and develop an education program with and for community optometrists regarding delivering falls prevention care for community-dwelling older adults<br>2. Gain practicing community optometrists' reaction to the education program | Page 7-8<br>See objectives |
| 2: Methods<br>*Provide a clear description of the methods used for PPI in the study* | Preparation: The education program was informed by prior work with community optometrists through a focus group.(2)<br><br>Phase 1: The program was designed by four community optometrists, three who were practicing regularly in clinical settings and one with expertise in curriculum design for university optometry programs. Two physiotherapists with experience in falls prevention education and community falls prevention also contributed to the design and development of the education program. They were included in the authorship team.<br><br>Phase 2: Participants were community optometrists and completed an online survey at baseline and post-education that included open-ended questions on their reaction and learnings from the falls prevention education program, as well as perceived barriers and enablers to implementing falls prevention care. Their feedback informed the assessment of acceptability and guided planned refinements to the program. | Page 11-12<br>See Phase 1: design of the Education<br><br>Pages 12-13<br>See Phase 2: Evaluation of the Education and Phase 2: Data Collection and Procedure |
| 3: Study Results<br>*Outcomes – Report the results of PPI in the study, including both positive and negative outcomes* | Community optometrists reported that the education program was acceptable, relevant and useful for clarifying their role in falls prevention. Their feedback highlighted perceived enablers such as concise clinical resources and opportunities for further training, as well as barriers such as time constraints, unclear referral pathways, and patient level barriers.<br><br>These insights will inform the assessment of program acceptability and guided planned refinements to support implementation of falls prevention care in community optometry settings. | Pages 16-26<br>See Results<br><br>Page 17-19<br>See Reaction to the education |
| 4: Discussion and conclusions<br>*Outcomes – Comment on the* | **Outcomes**<br>• Community optometrists' feedback confirmed that the structure and delivery of the education aligned with their expectations. Their | Pages 27-31<br>See Discussion |

| | | |
|---|---|---|
| *extent to which PPI influenced the study overall. Describe positive and negative effects* | involvement helped identify which design elements supported engagement and made the program easy to follow.<br>• Participants highlighted which component most effectively supported their learning and where additional clarification or emphasis was needed. Their reflections provided important context for interpreting how the education strengthened knowledge, awareness and confidence for falls prevention care.<br>• Participants identified perceived barriers and enablers that shaped their understanding of how falls prevention care could be incorporated into clinical practice. Their suggestions will inform considerations for refining the education to better support implementation. | |
| 5: Reflections/critical perspective *Comment critically on the study, reflecting on the things that went well and those that did not, so others can learn from this experience* | • The pilot study successfully developed and evaluated a falls education program for community optometrists. It was vital to engage with community optometrists because they were able to provide rich feedback during the design of the education regarding the clinical appropriateness and in phase 2, evaluate the education program for its relevance and suitability for their clinical practice.<br>• This study used pre/post surveys to evaluate changes in participants' knowledge, awareness and confidence comparing baseline and after viewing the education. The program was new and further studies with larger groups of optometrists will be needed to confirm its effectiveness.<br>• Optometrists offered valuable suggestions for improvements which will inform future program refinements. | Pages 30-31 See Strengths and Limitations |

Note: PPI = Patient and Public Involvement

Checklist available from Staniszewska S, Brett J, Simera I, Seers K, Mockford C, Goodlad S, et al. GRIPP2 reporting checklists: tools to improve reporting of patient and public involvement in research. BMJ (Online). 2017 Aug 2;358. doi.org/10.1136/bmj.j3453 (3)

S. Table 4. Pre/Post Education Survey

**(Where Likert Scales are used, response = strongly disagree, disagree, undecided, agree, and strongly agree**)

**Pre-Intervention Survey**

| | |
|---|---|
| 1. Have you come across falls prevention guidelines in optometry practice? | **Dichotomous** |
| 2. Are you aware that resources are available for optometrists to support falls prevention management? - awareness | **Dichotomous** |
| 3. What is the economic impact of falls in Australia (per annum)? **a) $4.7 billion b) $4.7 million c) $10.7 billion d) $1.7 billion** | **Multiple Choice** |
| 4. What is the prevalence of falls amongst older adults living in the community? **a) 1 in 3 b) 1 in 4 c) 1 in 5 d) 1 in 10 - knowledge** | **Multiple Choice** |
| 5. I feel I have the necessary knowledge to manage an older adult's falls risk in the clinic | **Likert Scale** |
| 6. I feel confident in routinely providing appropriate falls prevention management for older adults - confidence | **Likert Scale** |
| 7. I think falls prevention is relevant to my clinical practice - attitudes | **Likert Scale** |
| 8. I believe falls prevention education is important for improving patient outcomes - attitudes | **Likert Scale** |
| 9. I am open to incorporating falls prevention management into my current practices if it improves patient outcomes - motivation | **Likert Scale** |
| 10. In clinical practice, I can identify visual risk factors that increase a patient's risk of falls - awareness | **Likert Scale** |
| 11. I know what ocular diseases affect an older adult's risk of falling | **Likert Scale** |
| 12. I know the relevant referral pathways for older adults to receive falls prevention services if needed - confidence | **Likert Scale** |
| 13. I am willing to initiate referral to other allied health professionals to access multidisciplinary care - motivation | **Likert Scale** |
| 14. I believe optometrists should be embedded into a multidisciplinary team collaboration for falls prevention | **Likert Scale** |
| 15. Name up to **4** key ocular assessments required to understand an older adult's falls risk? - attitudes | **Open-Ended** |
| 16. Name up to **5** simple strategies optometrists can incorporate in their clinical practice to reduce older adults' falls risk | **Open-Ended** |
| 17. List **2** screening questions an optometrist can ask in clinic to understand an older adult's risk of falls | **Open-Ended** |
| 18. Describe how an optometrist can manage glasses and prescriptions for older adults to help prevent falls? | **Open-Ended** |

**Post-Intervention Survey**

| **First, we would like to explore your thoughts and opinions on the education program** | |
|---|---|
| 1. The education was easily accessible following the email instructions | **Likert-Scale** |
| 2. How long did it take you to complete the education programme? **In minutes** | **Insert number** |
| 3. The education programme makes its purpose completely evident **Disagree or Agree** | **Dichotomous** |
| 4. The programme presents information in a logical sequence **Disagree or Agree** | **Dichotomous** |

| | |
|---|---|
| 5. Did you feel the education programme was of an appropriate length?<br>**Too short, Acceptable, Too Long** | **Multiple Choice** |
| 6. Overall, the falls prevention education programme presented met my needs? | **Likert-Scale** |
| 7. The education programme used an appropriate range of media to demonstrate the key information | **Likert Scale** |
| 8. The short video at the beginning of the education contained enough information | **Likert-Scale** |
| **We have some questions regarding the format and presentation of the education program** | |
| 9. Text on the screen is easy to read<br>**Disagree or Agree** | **Dichotomous** |
| 10. The programme included information in breaks or "chunks"<br>**Disagree or Agree** | **Dichotomous** |
| 11. The programme uses visual cues (e.g. arrows, boxes, larger fonts) to draw attention to key points<br>**Disagree or Agree** | **Dichotomous** |
| 12. The programme sections have informative headers<br>**Disagree or Agree** | **Dichotomous** |
| 13. The programme provides a summary statement<br>**Disagree or Agree** | **Dichotomous** |
| 14. The programme uses illustrations and photographs that are clear and uncluttered<br>**Disagree or Agree** | **Dichotomous** |
| 15. The programme uses simple tables with short and clear row and column headings<br>**Disagree or Agree** | **Dichotomous** |
| 16. Did the education provide clear instructions on how optometrists can incorporate falls prevention into their clinical examination?<br>**Disagree or Agree** | **Dichotomous** |
| 17. The programme breaks down any action into manageable, explicit steps<br>**Disagree or Agree** | **Dichotomous** |
| **The following question will target the knowledge gained from the education program** | |
| 18. Have you come across falls prevention guidelines in optometry practice? | **Dichotomous** |
| 19. Are you aware that resources are available for optometrists to support falls prevention management? | **Dichotomous** |
| 20. What is the economic impact of falls in Australia (per annum)?<br>**a) $4.7 billion b) $4.7 million c) $10.7 billion d) $1.7 billion** | **Multiple Choice** |
| 21. What is the prevalence of falls amongst older adults living in the community?<br>**a) 1 in 3 b) 1 in 4 c) 1 in 5 d) 1 in 10** | **Multiple Choice** |
| 22. I feel I have the necessary knowledge to manage an older adult's falls risk in clinic | **Likert Scale** |
| 23. I feel confident in routinely providing appropriate falls prevention management for older adults | **Likert Scale** |
| 24. I think falls prevention is relevant to my clinical practice | **Likert Scale** |
| 25. I believe falls prevention education is important for improving patient outcomes | **Likert Scale** |
| 26. I am open to incorporating falls prevention management into my current practices if it improves patient outcomes | **Likert Scale** |
| 27. In clinical practice, I can identify visual risk factors that increase a patient's risk of falls | **Likert Scale** |
| 28. I know what ocular diseases affect an older adult's risk of falling | **Likert Scale** |

| | |
|---|---|
| 29. I know the relevant referral pathways for older adults to receive falls prevention services if needed | **Likert Scale** |
| 30. I am willing to initiate referral to other allied health professionals to access multidisciplinary care | **Likert Scale** |
| 31. I believe optometrists should be embedded into a multidisciplinary team collaboration for falls prevention | **Likert Scale** |
| 32. Name up to **4** key ocular assessments required to understand an older adult's falls risk? | **Open-Ended** |
| 33. Name up to **5** simple strategies optometrists can incorporate in their clinical practice to reduce older adults' falls risk | **Open-Ended** |
| 34. List **2** screening questions an optometrist can ask in clinic to understand an older adult's risk of falls | **Open-Ended** |
| 35. Describe how an optometrist can manage glasses and prescriptions for older adults to help prevent falls? | **Open-Ended** |
| **Please provide any suggestions for improving the education program** | |
| 36. How has the education changed your confidence to manage older adult's falls risk? | **Open-Ended** |
| 37. What barriers might challenge the implementation of falls prevention management into your practice? | **Open-Ended** |
| 38. What enablers might facilitate the implementation of falls prevention management into your practice? | **Open-Ended** |
| 39. What suggestions do you have to help us improve the education programme for future iterations? | **Open-Ended** |
| 40. Any other comments you would like to share regarding the education? | **Open-Ended** |

S.Table 5. Coding matrix of open ended responses with exemplar quotes

| Codes | Category | Exemplar Quote |
|---|---|---|
| Knowledge<br>Awareness<br>Resources<br>Guidelines | Knowledge and awareness of falls prevention for optometrists | "Some of the things these were outlined in the module were completely new to me"(P9).<br><br>"I am more informed on the general [falls] guidelines and resources available to educate myself and my patients" (P3)<br><br>"[The education] made me more aware of what to ask patients, and now I know where to find resources" (P4) |
| Implication<br>Manage<br>Role<br>Encourage | Changes in attitudes regarding enacting falls prevention care in clinical practice | "I have learnt about the implications [of falls risk] and I am more motivated to manage fall risk" (P6)<br><br>"Yes, it has reminded me that we play a useful role in [falls prevention]" (P13). |
| Confidence<br>Statistics<br>Advice | Changes in confidence regarding enacting falls prevention into practice | "I now feel more knowledgeable and confident in my ability to screen for falls risk and be able to take actionable steps to reduce the risk of falls in my patients" (P1).<br><br>"The education has improved the overall confidence for me to undertake falls prevention management" (P8). |
| Easy to digest<br>Practical advice<br>Sequence<br>Illustrations | Reaction to the education | "The advice on lighting conditions and the discussion around early cataract surgery has been very helpful with how I will discuss cataracts with my patients and encouraging patients not to delay cataract surgery" (P7).<br><br>"The CPD was simple to digest and broken down into dot points that are easy to remember for optometrists to incorporate falls prevention in practice" (P2).<br><br>"Some of the sizing of the visuals are a bit small, making the words hard to read without zooming in and out" (P2).<br><br>"Helpful to have "clinic ready" info eg. guidelines for what to put in a handout" (P11). |
| Time Constraints<br>Cost of Services<br>Patient understanding<br>Lack of equipment<br>Location of practice<br>Access to services | Barriers to incorporating falls prevention care into practice | "Optometrists working in retail practices (20-30 min appts) there may not be enough time to ask all questions and do all the appropriate tests" (P4).<br><br>"Patient lack of awareness and fear of losing independence" (P3). |
| Infographics<br>Pamphlets<br>Education material | Enablers to incorporating falls prevention care into practice | "A page printout or webpage that can be saved as a tab on the computer in dot point guideline form would be really good to have as a reference to refer to during consult" (P2). |

| | | |
|---|---|---|
| Collaboration | | "Clinical scenarios or case studies to simulate practical scenarios" (P3).<br><br>"An infographic printout so we could give to our patients would be nice" (P5). |

S.Table 6. Pre/Post Education responses to clinical knowledge about falls prevention care

| **Pre Education Survey** | |
|---|---|
| **Ocular assessment** | 1. Contrast sensitivity, visual acuity, stereopsis, ocular health exam (anterior + posterior eye) |
| | 2. Visual acuity, visual field, ocular health assessment |
| | 3. Visual field, Visual acuity, Stereoacuity, Contrast sensitivity |
| | 4. Visual acuity, visual field |
| | 5. Visual acuity, depth perception/ stereo, contrast sensitivity, refraction |
| | 6. Visual fields, Visual acuity, Contrast sensitivity |
| | 7. Visual acuity, visual fields, motility, contrast sensitivity |
| | 8. Visual acuity, contrast sensitivity, visual function/field and posterior eye health |
| | 9. Visual acuity, visual fields, anterior SLE and fundoscopy |
| | 10. Visual acuity, Visual Field, Slit Lamp, DFE |
| | 11. Visual acuity, visual field, contrast sensitivity; eye health assessment (eg. Slit lamp for cataract, dilated fundus examination for macular degeneration/glaucoma) |
| | 12. Visual acuity, visual field, contrast sensitivity, motility |
| | 13. Contrast sensitivity, visual fields, are they binocular, do they wear glasses |
| **Simple strategies** | 1. asking about previous falls, discussing living setting (stairs, ramps), in home aids (railings), discuss importance of lighting the environment |
| | 2. Raise awareness, Patient education on prevention |
| | 3. Risk assessment - history taking, ocular assessment, Refer to general practitioners for occupational therapists home assessment, Low vision referrals for relevant patients, |
| | 4. talk to patients about glasses adaptation, careful with lens type you prescribe |
| | 5. Advise regular eye exams, recommend single vision distance glasses, or ask patients to remove bifocals/multifocals when walk up and down the stairs (if applicable), trial framing to demo change in prescriptions, advise of adaptation period, refer for visual aids for patient with visual impairments/ low vision |
| | 6. History taking, discussion on multifocal adaptation |
| | 7. Ask about prior falls or balance issues, avoid putting older patients into multifocals, correct distance vision |
| | 8. Smaller prescription changes for elderly, recommending single vision lenses where appropriate especially for walking around, identify key eye conditions like macular degeneration and glaucoma, improving lighting in homes, advising an occupational therapist where needed, advise low vision services like orientation mobility training etc. |
| | 9. Multi focal lenses education, visuals field confrontation's screening, good history taking, dilated funds examinations on indication, communication with general practitioners |
| | 10. Handles/bars to help patients get up and sit down, bright lighting, eliminating steps, |
| | 11. Ensure appropriate correction; consider using single vision distance glasses instead of bifocals/multifocals for walking around; encourage highlighting of steps with yellow tape to improve visibility; setting bifocals/mutlifocals lower to reduce risk; improving lighting at home/areas where falls are likely. |
| | 12. actively ask about falls, observe patient gait and behaviour in clinic, communication with general practitioners and other health providers |

| | |
|---|---|
| | 13. no mats, bright contrast between floor and step up to seat, get step out of way for them to climb on, allow them to bring walker into room. |
| **Screening question** | 1. Have you had any falls? Do you feel unsteady at times? |
| | 2. Have you had any recent injury in which you feel like it may have been related to your vision |
| | 3. History of falls, dizziness/imbalance when walking |
| | 4. Have you had a fall before, do you have mobility issues |
| | 5. Any trouble with balance, tripping or falling? Can you see clearly at distance with glasses / without glasses? |
| | 6. Do you feel uneasy or lack confidence when on your own (travelling), how many near misses or actual falls have you had in the last 6 months. |
| | 7. Have you had any previous falls/how is your balance? |
| | 8. Do you navigate unfamiliar environments with confidence? Are there any issues with balance, the hips or the legs? |
| | 9. "How do you go with navigating your environment day to day?", "have you experienced and falls or injuries recently" |
| | 10. Have you had any fall recently? Do you feel that your vision impacts how you judge steps or uneven ground |
| | 11. Have you had any falls recently?; do you find yourself feeling unsteady or uncertain if in unfamiliar areas or places where the ground is irregular? |
| | 12. Have you ever had a fall, have you ever had a "almost fall" |
| | 13. Do you exercise regularly, how is your balance |
| **Optical management** | 1. Maximising binocular vision, prioritising depth perception and clarity over convenience (e.g. multifocals in elderly populations should be prescribed with caution), explaining changes to stereopsis with sudden visual loss or degenerative eye disease, tints to improve colour vision discrimination |
| | 2. Patient education? |
| | 3. Avoid large change in refractive correction , avoid multifocals lenses to older single-vision lens wearers |
| | 4. Discuss adaptation, don't prescribe large jumps in prescription, carefully consider lens type to avoid prism jump/warping or distortion |
| | 5. Reduce prescription if there's been a large change, warn of how bifocals/multifocals work for first time wearer, encourage slowly introduce new bifocals/multifocals into daily life, promote single vision distance glasses full time wear if it improves their vision, prisms if diplopia is present |
| | 6. Ensure appropriate frame size, prisms if needed. |
| | 7. Avoid multifocals, optimise distance vision |
| | 8. To prescribe minimal changes: less than or equal to 0.50 changes in both sphere and cylinder and axis changes of less than 15 degrees. To provide glasses to individuals with visual acuity less than 6/12 unaided as this can signficantly affect falls risks. Providing single vision lenses to patients with ocular conditions which affect peripheral or central vision. |
| | 9. Multifocals/bifocals fitted to ensure comfort when looking down otherwise having a single vision distance/reading glasses combination so the patient doesn't have to deal with peripheral distortion or increased magnification in the lower portion of the lenses |
| | 10. Give them the best vision possible, advise not to use multifocal spectacles to walk up and down any stairs if they have never tried them before. |
| | 11. Using distance glasses only prescription for walking around; being mindful of changes in space perception with sudden changes in prescription (eg. Myopic |

| | |
|---|---|
| | shift due to cataract); advice to ophthalmology re single vision being a better option post-cataract surgery vs monovision to reduce risk of falls; using tints judiciously to assist with glare where necessary |
| | 12. Choose appropriate size frames, avoid multifocal/bifocals in new elderly wearers (consider if existing multifocal/bifocals are working well for elderly patients) |
| | 13. Make sure they are up to date and wearing the correct pair for task |
| **Post Education Responses** | |
| **Ocular assessment** | 1. Visual acuity, contrast sensitivity, visual fields, stereoacuity |
| | 2. Visual acuity, contrast sensitivity, visual field, stereoacuity |
| | 3. Visual acuity, visual field, melbourne contrast sensitivity, refraction |
| | 4. Visual acuity, visual field, contrast sensitivity, stereo |
| | 5. Visual acuity, stereoacuity, ocular health check, visual field |
| | 6. Visual acuity, visual fields, fundus exam |
| | 7. Visual acuity, refraction, contrast sensitivity, visual fields |
| | 8. Visual acuity, contrast sensitivity, visual field, refractive error |
| | 9. Visual acuity, contrast sensitivity, visual fields, stereoacuity |
| | 10. Visual acuity, contrast sensitivity, visual field, depth perception |
| | 11. Visual acuity, visual contrast, visual field, stereopsis |
| | 12. Acuity, contrast sensitivity, fields, refractive error |
| | 13. Visual acuity, visual field, contrast sensitivity, depth perception |
| **Simple strategies** | 1. Screening, ocular assessment, prescribe visual aids, offer advice, referral |
| | 2. Screening, relevant ocular assessment, prescribing appropriate visual aids, referral to multidisciplinary team for high risk patients, provide brief advice on falls prevention |
| | 3. Single vision glasses for uncorrected hyperopes, history taking: screening for risk factors of falls, referring for early cataract surgery, ensure optimum refractive correction for easy adaptation , refer to general practitioner to initiate multidisciplinary approach for fall prevention, general advice to reduce fall risk ( ensure proper lighting and footwear) |
| | 4. Ask questions, don't change prescription too much, do appropriate clinical tests, advise patient about lighting and environmental factors, avoid multifocal/bifocals |
| | 5. Screening risks, proper eye exam, prescribe appropriate visual aids (single vision over multifocals), brief advice re: what can be done to reduce fall risks, and referral to appropriate health professionals if need |
| | 6. Observe gait/disability/weaknesses, ask questions during history |
| | 7. Consider single vision glasses instead of bifocals/multifocal glasses, ask in history about falls/risks, correct distance refractive error, discuss falls risks with patients, initiate referrals to other practitioners involved in care |
| | 8. Advice on lighting and home safety assessment, screening for falls history and causes, ocular health assessment and guiding based on visual function loss. E.g. Careful of tripping over things with inferior arcuate defects, giving them the right glasses for what they need, managing current ocular health conditions, e.g. macular degeneration, glaucoma, cataracts, diabetes. Referring to appropriate allied health professional services. |

| | |
|---|---|
| | 9. Screening during history taking, ocular assessments, prescribing appropriate visual aids, offer brief advice for falls, referral pathways to other allied health professionals |
| | 10. Screening, ocular assessments, prescribe visual aids, refer |
| | 11. Refer for assessment of lighting etc at home, recommend exercise regime with physio/general practitioner; refer for early cataract surgery; minimise sudden changes in prescription; correct hyperopia even if visual acuity ok |
| | 12. Assessment of contrast sensitivity, consider early cataract referral, sensible glasses prescribing choices, actively asking regarding falls |
| | 13. Make sure they have glasses that are correct, don't change the prescriptions too dramatically all at once, check their contrast sensitivity, recommend the use of single vision glasses when outside or walking. |
| **Screening question** | 1. Have you had any falls in the last year? If so, how frequent? |
| | 2. History of falls, mobility experience |
| | 3. Have you had any falls in the last year, have you had any issues with balance or dizziness when walking around |
| | 4. Have you fall before, do you ever feel dizzy |
| | 5. Have you had any falls this year? What was the context, severity and sequelae of the falls? |
| | 6. Frequency of falls in past 2 months, subjective difficulties such as finding it hard to see curbs |
| | 7. Have you had any previous falls? Do you ever feel dizzy or experience lack of balance? |
| | 8. Have you had any falls? How is your mobility? Do you feel confident and steady on your feet, if not, what have you struggled with when walking around? |
| | 9. Have you had any previous falls in the last year? Any concerns about falls or limited mobility? |
| | 10. Have you had any falls in last year? If so, how often? |
| | 11. Have you fallen in the last year; how often have you fallen? |
| | 12. Have you had a fall in the last year, have you had any near misses |
| | 13. Have you had a fall in the past year? Do you have trouble getting out of a chair |
| **Optical management** | 1. Limit large changes in prescriptions, educate regarding depth perception changes and image shifts with bifocals/multifocals |
| | 2. Prescribe appropriate visual aids and avoid unnecessary prescription of bifocals or multifocals, and provide advice on appropriate use of glasses prescribed |
| | 3. Single vision glasses for uncorrected hyperopia and single vision distance glasses for elderly use, conservative prescription to avoid large changes |
| | 4. Avoid bifocals/multifocals, avoid changing prescriptions too much |
| | 5. Avoid prescribing large changes all at once. Single vision over multifocals for older adults especially for walking purposes. Correct hyperopia |
| | 6. Ensure glasses have good field of view and fit well. |
| | 7. Prescribing distance refractive error, opting for single vision distance over bifocals/multifocals for patients at risk of falls |
| | 8. Prescribing single vision lenses |

| | |
|---|---|
| | 9. Single vision distance specs for every day navigation, minimal prescription jumps if they are a previous prescription wearer. Good for patients with uncorrected hyperopia as well. |
| | 10. Glasses improves patients' visual acuity |
| | 11. Use single vision distance glasses instead of multifocal/bifocals; avoid changing prescription by more than 0.75; avoid monovision; correct hyperopia |
| | 12. Avoid large changes to refraction, avoid multifocals/bifocals in elderly patients who have not worn them before, for existing bifocals/multifocals wearers consider single vision distance specs for use outside the home |
| | 13. Go the single vision distance glasses for outside and walking, and make sure those uncorrected high hyperopes get some glasses to see properly with even if they "don't need them". |
